# The Impact of Malnutrition Severity on Mortality Risk of Older Adults with Acute Heart Failure: A Geriatric Nutritional Risk Index-Based Assessment

**DOI:** 10.1101/2025.09.29.25336938

**Authors:** Tsukasa Murakami, Keisuke Kojima, Masanori Takenoya, Kentaro Jujo, Ryusuke Ae, Masanari Kuwabara

**Affiliations:** Department of Cardiology, Japanese Red Cross Ogawa Hospital, 1525 Ogawa, Ogawa-machi, Hiki, Saitama, Japan; Department of Cardiology, Saitama Medical Center, 1981 Kamoda, Kawagoe, Saitama, Japan; Division of Cardiology, Department of Medicine, Nihon University School of Medicine, 30-1 Ohyaguchi-kamicho, Itabashi-ku, Tokyo, Japan; Division of Public Health, Center for Community Medicine, Jichi Medical University, Tochigi, 329-0498, Japan; Division of Cardiovascular Medicine, Department of Medicine, Jichi Medical University, Tochigi, Japan

**Keywords:** Malnutrition, Geriatric Nutritional Risk Index, Acute heart failure, Older adults

## Abstract

**Backgrounds:** Malnutrition is associated with increased risk of all-cause mortality in patients with acute heart failure (AHF). However, the increasing mortality risk associated with the severity of malnutrition in older adults with AHF remained unclear. We aimed to investigate the prognostic impact of malnutrition severity on all-cause mortality of older adults with AHF.

**Methods:** This single-center study included older adults (aged ≥65) hospitalized for AHF between 2019 and 2023. We used the Geriatric Nutritional Risk Index (GNRI) as a proxy for nutritional status, dividing patients into the normal (GNRI>98, n=64), mild risk (GNRI 92-98, n=54), moderate risk (GNRI 82-<92, n=66), and severe risk (GNRI<82, n=30) groups. The study outcome was all-cause mortality.

**Results:** We included 214 patients (mean age, 85±8 years; male, 49%). During the median follow-up of 356 days (interquartile range, 66-919 days), 76 deaths were observed. In the patients’ background, worse GNRI was associated with older age, underweight, frailty, and anemia. Multivariable Cox hazard analysis revealed that moderate risk (HR, 2.69; 95%CI, 1.34-5.40; p=0.01) and severe risk (HR, 9.75; 95% CI, 4.30-22.10; p<0.01) were associated with higher mortality compared with normal GNRI, in addition to aging (HR per 1 year increase, 1.07; 95%CI, 1.03-1.11; p<0.01). A sensitivity analysis with the continuous value of GNRI demonstrated that lower GNRI was associated with higher mortality (HR, 0.92; 95% CI, 0.90-0.95; p<0.01). The association of the GNRI with all-cause mortality was consistent in the subgroup analysis of age ≥85 years, sex, body mass index ≤18.5, frailty, and anemia.

**Conclusions:** The moderate and severe GNRI risk categories were associated with higher all-cause mortality. Malnutrition severity assessed by the GNRI could be useful in estimating the risk of older adults with AHF, even in the oldest-old (those aged 85 years or older).

## Introduction

As ageing accelerates, the number of patients with heart failure (HF) has increased significantly (1). Acute heart failure (AHF) is a critical condition with a 1-year mortality of 11-27% (2-6), especially for older adults (5, 6). To screen for high-risk patients requiring comprehensive care, the risk assessment for older adults with AHF is critically important early after admission.

Malnutrition is common among older adults with AHF and is associated with increased mortality (7-15). The Geriatric Nutritional Risk Index (GNRI) is an objective tool to assess the nutritional risk of older adults (16). Malnutrition assessed by the GNRI shows an association with high mortality of patients with AHF (8, 9, 12-15). However, many studies dichotomized the study population by defining malnutrition as GNRI <92 (8, 12-15) or <98 (9). Therefore, the increasing risk associated with the severity of malnutrition remained unclear. Furthermore, the impact of the GNRI-based nutritional assessment on the mortality of the oldest-old patients (those aged 85 years or older) with AHF has not been fully investigated. We hypothesized that worse nutritional status at admission could be associated with higher mortality and could be useful in estimating the risk for older adults with AHF. This study aimed to investigate the prognostic impact of malnutrition severity assessed by the GNRI on all-cause mortality of older adults with AHF.

## Methods

This was an observational, single-center study. We reviewed consecutive patients with AHF who were admitted to the Japanese Red Cross Ogawa Hospital (JRCOH) (Saitama, Japan) between January 2019 and December 2023. The JRCOH is a secondary emergency hospital in an ageing area, where approximately 40% of residents are aged 65 years or older. We screened emergency hospitalized patients with the diagnostic codes of HF in the ICD-10 (I50.1, I50.1, I50.9, I11.0, I13.0, and I13.2) (17). From them, patients aged <65 years old and patients whose GNRI could not be calculated at admission were excluded. We acquired clinical information from the hospital records. This study was approved by the institutional review board (approval number, Ogawa-Rin-90, March 12, 2025), and was performed in accordance with the Declaration of Helsinki. Since this was a retrospective study without further invasion, the opt-out method was used.

The GNRI is an objective tool created to predict malnutrition-related complications in older adults (16). The GNRI was calculated as follows: 14.89×serum albumin level (g/dl)+41.7×(body weight (kg) /ideal body weight (kg)) (16). When the body weight/ideal body weight ratio was greater than 1, the value was set to 1, as in the previous studies (16, 18). The ideal body weight was calculated by using the Lorentz formula as follows: ideal body weight for men (kg) = height (cm) − 100 − (height (cm) − 150)/4 and for women (kg) = height (cm) − 100 − (height (cm) − 150)/2.5 (16, 18). GNRI >98 was classified as normal, GNRI of 92 to 98 as mild nutritional risk, GNRI of 82 to <92 as moderate nutritional risk, and GNRI <82 as severe nutritional risk, respectively (16). Besides the GNRI, Controlling Nutritional Status (CONUT) scores and Prognostic Nutritional Index (PNI) were also calculated as other nutritional assessment tools (19, 20). The CONUT score was calculated by summing up designated points of three laboratory markers (serum albumin, total lymphocyte counts, and total cholesterol). CONUT score ranges from 0 to 12 (19). The PNI was calculated as follows: 10 × serum albumin in g/dL + 0.005 × total lymphocyte count in /μL (20). Frailty was evaluated using the Clinical Frailty Scale (CFS) (21). According to the CFS, patient frailty before admission was scored as follows: (i) very fit; (ii) well; (iii) managing well; (iv) vulnerable; (v) mildly frail; (vi) moderately frail; (vii) severely frail; (viii) very severely frail; and (ix) terminally ill (21). Using echocardiography, left ventricular ejection fraction (LVEF) and valvular diseases of the left heart were examined. Significant valve disease was defined as worse than moderate in severity. The primary outcome was all-cause mortality.

### Statistical analysis

Data are presented as percentages for categorical variables, mean ± standard deviation for normally distributed continuous variables, or a median and interquartile range for non-normally distributed continuous variables. Statistical significance was set at p < 0.05. The study population was divided into four GNRI categories: normal, mild risk, moderate risk, and severe risk based on their GNRI at admission. First, the baseline characteristic among the four groups was compared. Categorical variables were compared using the Fisher’s exact test. Normally distributed continuous variables were compared using one-way ANOVA. Otherwise, continuous variables were compared using the Kruskal–Wallis test. The cumulative incidence of all-cause death was compared using the Kaplan-Meier method. Then, we performed a multivariable Cox hazard analysis to calculate the hazard ratio (HR) of the GNRI for all-cause mortality. Considering the small number of events and missing data in BNP levels (missing in 14% of patients), we performed three multivariable analyses as follows. In model 1, the GNRI category was adjusted for age, sex, hypertension, diabetes mellitus, dyslipidemia, estimated glomerular filtration rate (GFR) <60 mL/min/1.73m², and New York Heart Association (NYHA) class. In model 2, prior HF admission, history of percutaneous coronary intervention (PCI) or coronary artery bypass grafting (CABG), cancer, and hemoglobin levels were further adjusted. In model 3, log BNP was additionally adjusted, and the other variables were the same as in model 2. To calculate the HR, the normal in the GNRI risk was used as a reference. As a sensitivity analysis, GNRI as a continuous metric was tested in these three models. Moreover, a subgroup analysis according to age, sex, body mass index (BMI), frailty, anemia, and presence of peripheral edema was performed. Patients with CFS ≥ 4 were defined as having frailty (21). Anemia was defined as a hemoglobin level <13.0 g/dL in men and <12.0 g/dL in women according to the criteria defined by the World Health Organization (22). In the subgroup analysis, the HR of the GNRI (as a continuous metric) for all-cause mortality was calculated after adjusting for age and sex. SPSS ver. 20 for Windows (SPSS, Inc., Chicago, Illinois) was used for the description of demographic data, survival analysis, and multivariable analysis. To draw a restricted spline curve, R v4.2.1 (R Foundation for Statistical Computing, Vienna, Austria) and packages of “rms”, “survival”, “dplyr”, and “ggplot2” were used.

## Results

During the study period, 374 patients were admitted on an emergently basis for AHF. Of them, 25 patients aged <65 years old and 135 patients whose GNRI could not be calculated (71 patients without recorded body weight, 31 patients without serum albumin levels, and 33 patients without both of these variables) at admission were excluded, resulting in 214 patients for the final study population (**Figure 1**). The mean age of the final study population was 85±8 years (male, 49%). They were divided into four GNRI risk groups: normal group (n=64, 30%), mild risk group (n=54, 25%), moderate risk group (n=66, 31%), and severe risk group (n=30, 14%). Patients with higher nutritional risks were older (82±8, 86±8, 86±7, and 85±8 years, in normal, mild risk, moderate risk, and severe risk groups, respectively, p=0.01), showed lower BMI (25.0±3.9, 23.7±3.2, 20.9±3.2, and 19.1±3.7, p<0.01), hemoglobin levels (12.4±2.3, 11.1±2.0, 10.7±2.5, and 10.8±1.8, p<0.01), and albumin levels (4.1±0.3, 3.6±0.2, 3.3±0.2, and 2.7±0.4, p<0.01), demonstrated higher CFS scores (2 [interquartile range, 2-4], 4 [2-5], 4 [2-5], and 7 [2-7], p<0.01), and had a higher prevalence of peripheral edema (64%, 78%, 82%, and 90%, p=0.03) (**Table 1**). Regarding the nutritional assessment tools, the CONUT and PNI showed consistent results with the GNRI.

**Figure 1.**
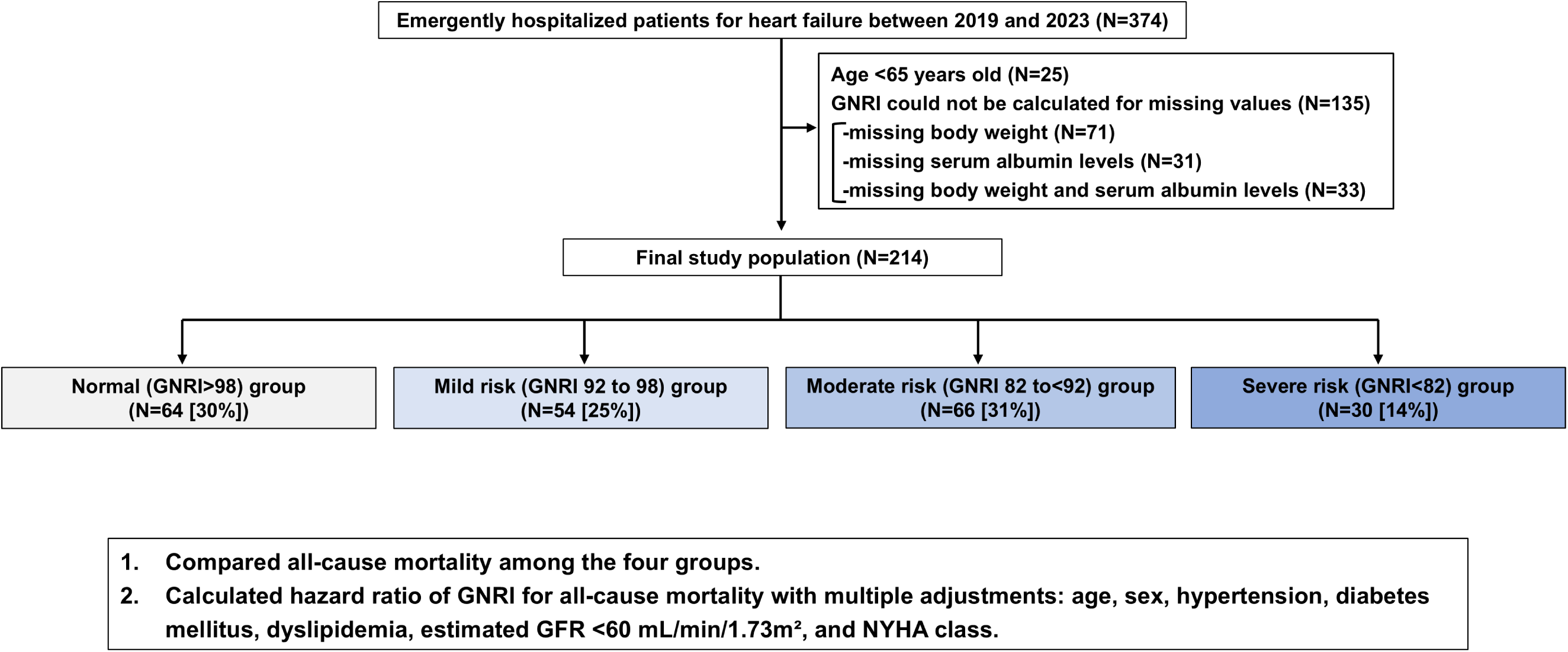
Study flow chart. Abbreviations: GFR, glomerular filtration rate; GNRI, Geriatric Nutritional Risk Index; NYHA, New York Heart Association.

**Table 1.**
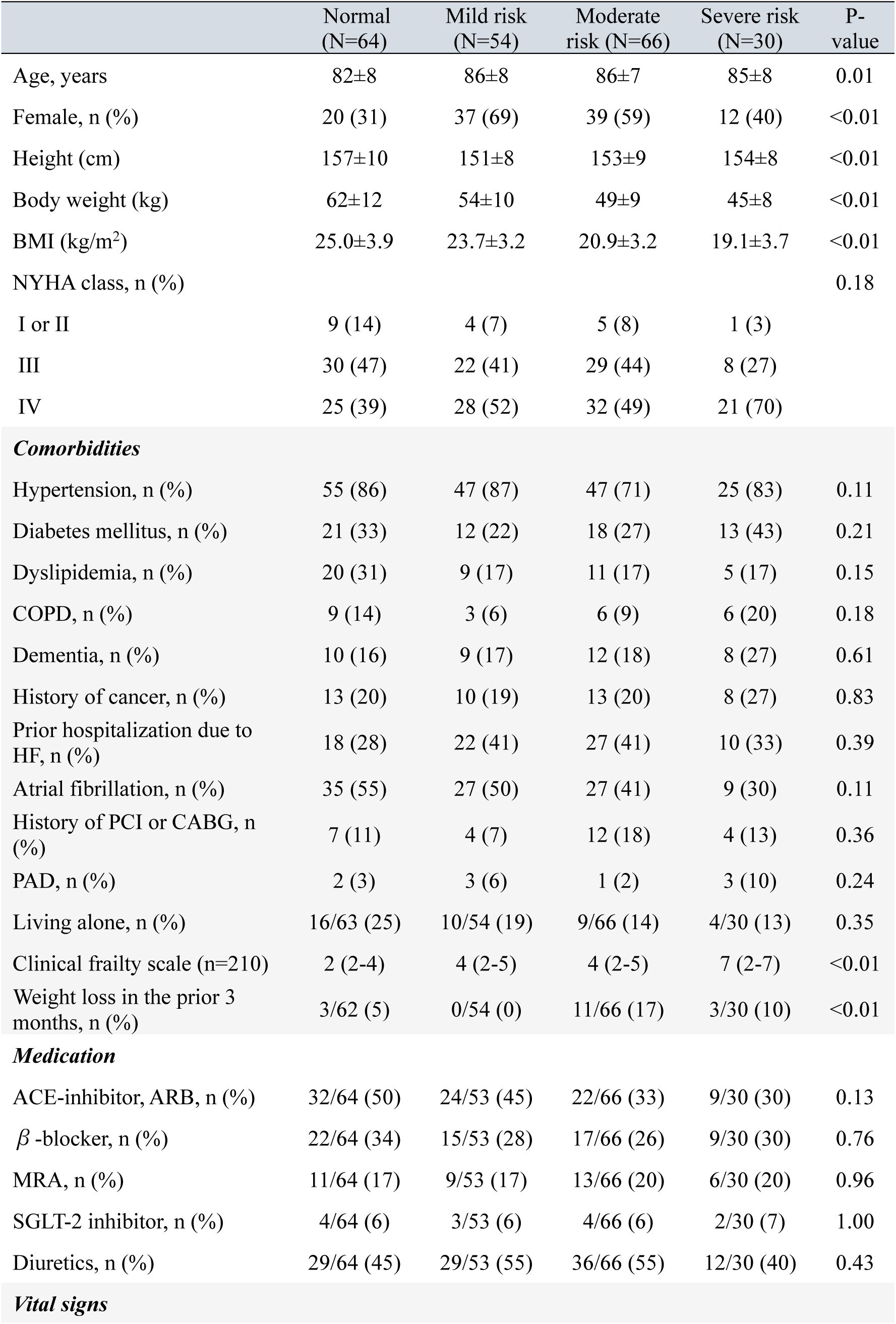
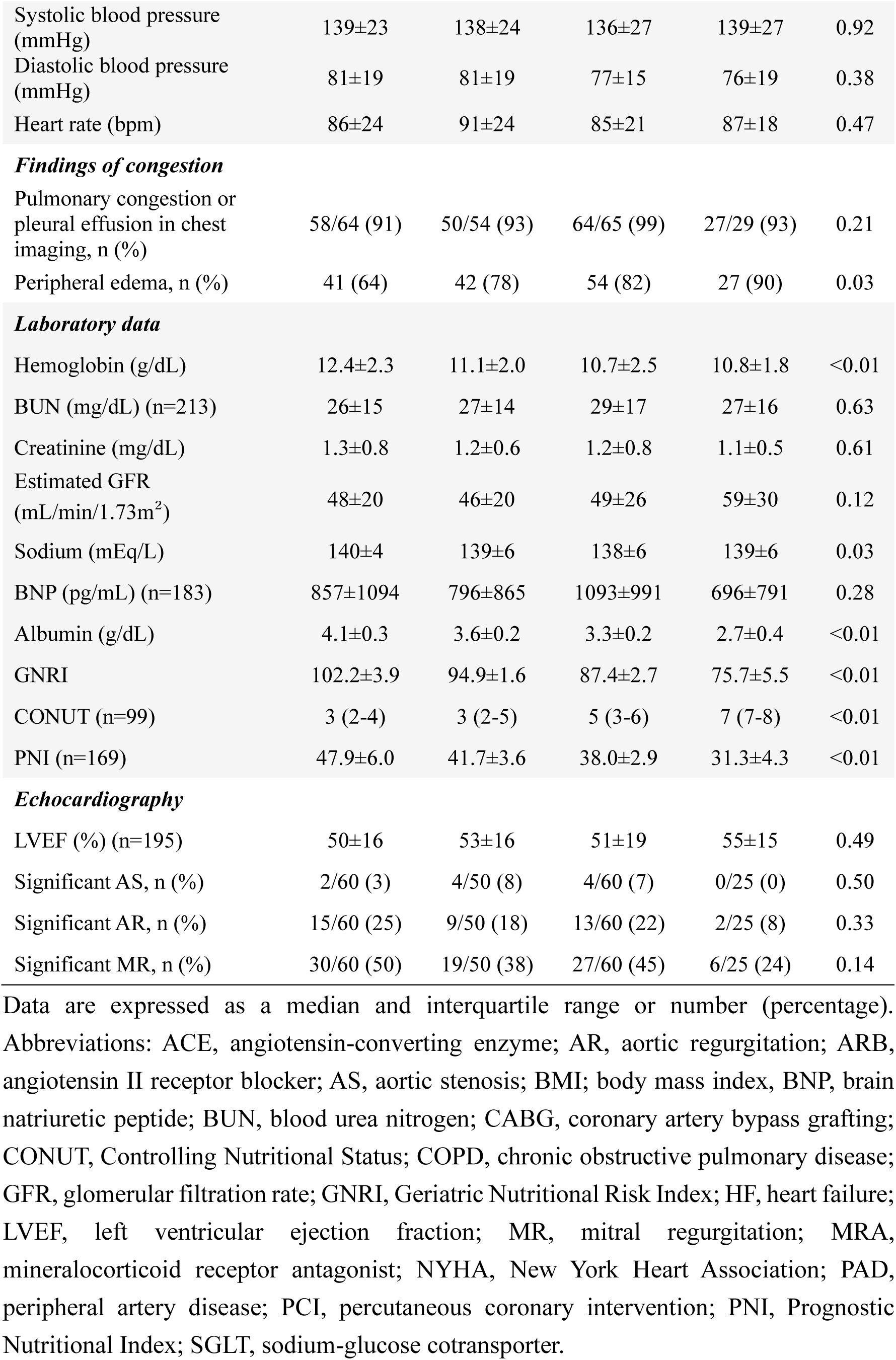
Clinical characteristics.

|  | Normal<br>(N=64) | Mild risk<br>(N=54) | Moderate<br>risk (N=66) | Severe risk<br>(N=30) | P-value |
| --- | --- | --- | --- | --- | --- |
| Age, years | 82±8 | 86±8 | 86±7 | 85±8 | 0.01 |
| Female, n (%) | 20 (31) | 37 (69) | 39 (59) | 12 (40) | <0.01 |
| Height (cm) | 157±10 | 151±8 | 153±9 | 154±8 | <0.01 |
| Body weight (kg) | 62±12 | 54±10 | 49±9 | 45±8 | <0.01 |
| BMI (kg/m<sup>2</sup>) | 25.0±3.9 | 23.7±3.2 | 20.9±3.2 | 19.1±3.7 | <0.01 |
| NYHA class, n (%) |  |  |  |  | 0.18 |
| I or II | 9 (14) | 4 (7) | 5 (8) | 1 (3) |  |
| III | 30 (47) | 22 (41) | 29 (44) | 8 (27) |  |
| IV | 25 (39) | 28 (52) | 32 (49) | 21 (70) |  |
| **Comorbidities** |  |  |  |  |  |
| Hypertension, n (%) | 55 (86) | 47 (87) | 47 (71) | 25 (83) | 0.11 |
| Diabetes mellitus, n (%) | 21 (33) | 12 (22) | 18 (27) | 13 (43) | 0.21 |
| Dyslipidemia, n (%) | 20 (31) | 9 (17) | 11 (17) | 5 (17) | 0.15 |
| COPD, n (%) | 9 (14) | 3 (6) | 6 (9) | 6 (20) | 0.18 |
| Dementia, n (%) | 10 (16) | 9 (17) | 12 (18) | 8 (27) | 0.61 |
| History of cancer, n (%) | 13 (20) | 10 (19) | 13 (20) | 8 (27) | 0.83 |
| Prior hospitalization due to HF, n (%) | 18 (28) | 22 (41) | 27 (41) | 10 (33) | 0.39 |
| Atrial fibrillation, n (%) | 35 (55) | 27 (50) | 27 (41) | 9 (30) | 0.11 |
| History of PCI or CABG, n (%) | 7 (11) | 4 (7) | 12 (18) | 4 (13) | 0.36 |
| PAD, n (%) | 2 (3) | 3 (6) | 1 (2) | 3 (10) | 0.24 |
| Living alone, n (%) | 16/63 (25) | 10/54 (19) | 9/66 (14) | 4/30 (13) | 0.35 |
| Clinical frailty scale (n=210) | 2 (2-4) | 4 (2-5) | 4 (2-5) | 7 (2-7) | <0.01 |
| Weight loss in the prior 3 months, n (%) | 3/62 (5) | 0/54 (0) | 11/66 (17) | 3/30 (10) | <0.01 |
| **Medication** |  |  |  |  |  |
| ACE-inhibitor, ARB, n (%) | 32/64 (50) | 24/53 (45) | 22/66 (33) | 9/30 (30) | 0.13 |
| β-blocker, n (%) | 22/64 (34) | 15/53 (28) | 17/66 (26) | 9/30 (30) | 0.76 |
| MRA, n (%) | 11/64 (17) | 9/53 (17) | 13/66 (20) | 6/30 (20) | 0.96 |
| SGLT-2 inhibitor, n (%) | 4/64 (6) | 3/53 (6) | 4/66 (6) | 2/30 (7) | 1.00 |
| Diuretics, n (%) | 29/64 (45) | 29/53 (55) | 36/66 (55) | 12/30 (40) | 0.43 |
| **Vital signs** |  |  |  |  |  |
| Systolic blood pressure (mmHg) | 139±23 | 138±24 | 136±27 | 139±27 | 0.92 |
| Diastolic blood pressure (mmHg) | 81±19 | 81±19 | 77±15 | 76±19 | 0.38 |
| Heart rate (bpm) | 86±24 | 91±24 | 85±21 | 87±18 | 0.47 |
| ***Findings of congestion*** |  |  |  |  |  |
| Pulmonary congestion or pleural effusion in chest imaging, n (%) | 58/64 (91) | 50/54 (93) | 64/65 (99) | 27/29 (93) | 0.21 |
| Peripheral edema, n (%) | 41 (64) | 42 (78) | 54 (82) | 27 (90) | 0.03 |
| ***Laboratory data*** |  |  |  |  |  |
| Hemoglobin (g/dL) | 12.4±2.3 | 11.1±2.0 | 10.7±2.5 | 10.8±1.8 | <0.01 |
| BUN (mg/dL) (n=213) | 26±15 | 27±14 | 29±17 | 27±16 | 0.63 |
| Creatinine (mg/dL) | 1.3±0.8 | 1.2±0.6 | 1.2±0.8 | 1.1±0.5 | 0.61 |
| Estimated GFR (mL/min/1.73m<sup>2</sup>) | 48±20 | 46±20 | 49±26 | 59±30 | 0.12 |
| Sodium (mEq/L) | 140±4 | 139±6 | 138±6 | 139±6 | 0.03 |
| BNP (pg/mL) (n=183) | 857±1094 | 796±865 | 1093±991 | 696±791 | 0.28 |
| Albumin (g/dL) | 4.1±0.3 | 3.6±0.2 | 3.3±0.2 | 2.7±0.4 | <0.01 |
| GNRI | 102.2±3.9 | 94.9±1.6 | 87.4±2.7 | 75.7±5.5 | <0.01 |
| CONUT (n=99) | 3 (2-4) | 3 (2-5) | 5 (3-6) | 7 (7-8) | <0.01 |
| PNI (n=169) | 47.9±6.0 | 41.7±3.6 | 38.0±2.9 | 31.3±4.3 | <0.01 |
| ***Echocardiography*** |  |  |  |  |  |
| LVEF (%) (n=195) | 50±16 | 53±16 | 51±19 | 55±15 | 0.49 |
| Significant AS, n (%) | 2/60 (3) | 4/50 (8) | 4/60 (7) | 0/25 (0) | 0.50 |
| Significant AR, n (%) | 15/60 (25) | 9/50 (18) | 13/60 (22) | 2/25 (8) | 0.33 |
| Significant MR, n (%) | 30/60 (50) | 19/50 (38) | 27/60 (45) | 6/25 (24) | 0.14 |

Data are expressed as a median and interquartile range or number (percentage). Abbreviations: ACE, angiotensin-converting enzyme; AR, aortic regurgitation; ARB, angiotensin II receptor blocker; AS, aortic stenosis; BMI, body mass index, BNP, brain natriuretic peptide; BUN, blood urea nitrogen; CABG, coronary artery bypass grafting; CONUT, Controlling Nutritional Status; COPD, chronic obstructive pulmonary disease; GFR, glomerular filtration rate; GNRI, Geriatric Nutritional Risk Index; HF, heart failure; LVEF, left ventricular ejection fraction; MR, mitral regurgitation; MRA, mineralocorticoid receptor antagonist; NYHA, New York Heart Association; PAD, peripheral artery disease; PCI, percutaneous coronary intervention; PNI, Prognostic Nutritional Index; SGLT, sodium-glucose cotransporter.

During the median follow-up of 356 days (interquartile range, 66-919 days), 76 all-cause deaths were observed. The cumulative incidence of all-cause mortality was the highest in the severe risk group (53%), followed by the moderate risk group (36%), the mild risk group (33%), and the normal group (28%) (p<0.01) (**Figure 2**). Similarly, cumulative 1-year mortality was the highest in the severe risk group (50%), followed by the moderate risk group (30%), the mild risk group (13%), and the normal group (9%) (p<0.01). After adjustments for age and sex, a restricted spline curve demonstrated the association between lower GNRI (continuous metric) and a higher risk of all-cause mortality (p for overall association <0.01, p for non-linearity = 0.18) (**Figure 3**). Multivariable Cox hazard analysis revealed that moderate risk (HR, 2.69; 95% CI 1.34-5.40; p = 0.01) and severe risk (HR, 9.75; 95% CI 4.30-22.10; p<0.01), and age (HR per 1 increment, 1.07; 95% CI 1.03-1.11; p<0.01) were associated with all-cause mortality after multiple adjustments in model 1 (**Table 2**). Mild risk (HR, 1.14; 95% CI 0.57-2.29; p = 0.71) was not significantly different from normal GNRI. In model 2, moderate risk (HR, 2.61; 95% CI 1.26-5.38; p=0.01) and severe risk (HR, 9.62; 95% CI 4.20-22.02; p<0.01) remained significant after further adjustments. However, in model 3, severe risk (HR, 8.12; 95% CI 3.07-21.48; p<0.01) remained statistically significant after adjusting for log BNP; conversely, moderate risk was not statistically significant (**Supplementary Table 1**). The sensitivity analysis, in which GNRI was treated as a continuous variable, revealed that lower GNRI was associated with higher all-cause mortality after multiple adjustments as in model 1 (HR per 1 increment, 0.92; 95% CI 0.90-0.95; p<0.01) (**Table 2**). In addition, the GNRI in a continuous metric was consistently associated with all-cause mortality after further adjustments, as in models 2 and 3 (**Supplementary Table 1**). Moreover, the GNRI (as a continuous metric) showed a consistent association with all-cause mortality, regardless of age ≥85, sex, BMI <18.5, CFS ≥4, and anemia (**Figure 4**). The GNRI was also associated with all-cause mortality in patients with edema. Although not statistically significant, a numerical tendency for higher mortality with lower GNRI was observed in patients without edema.

**Figure 2.**
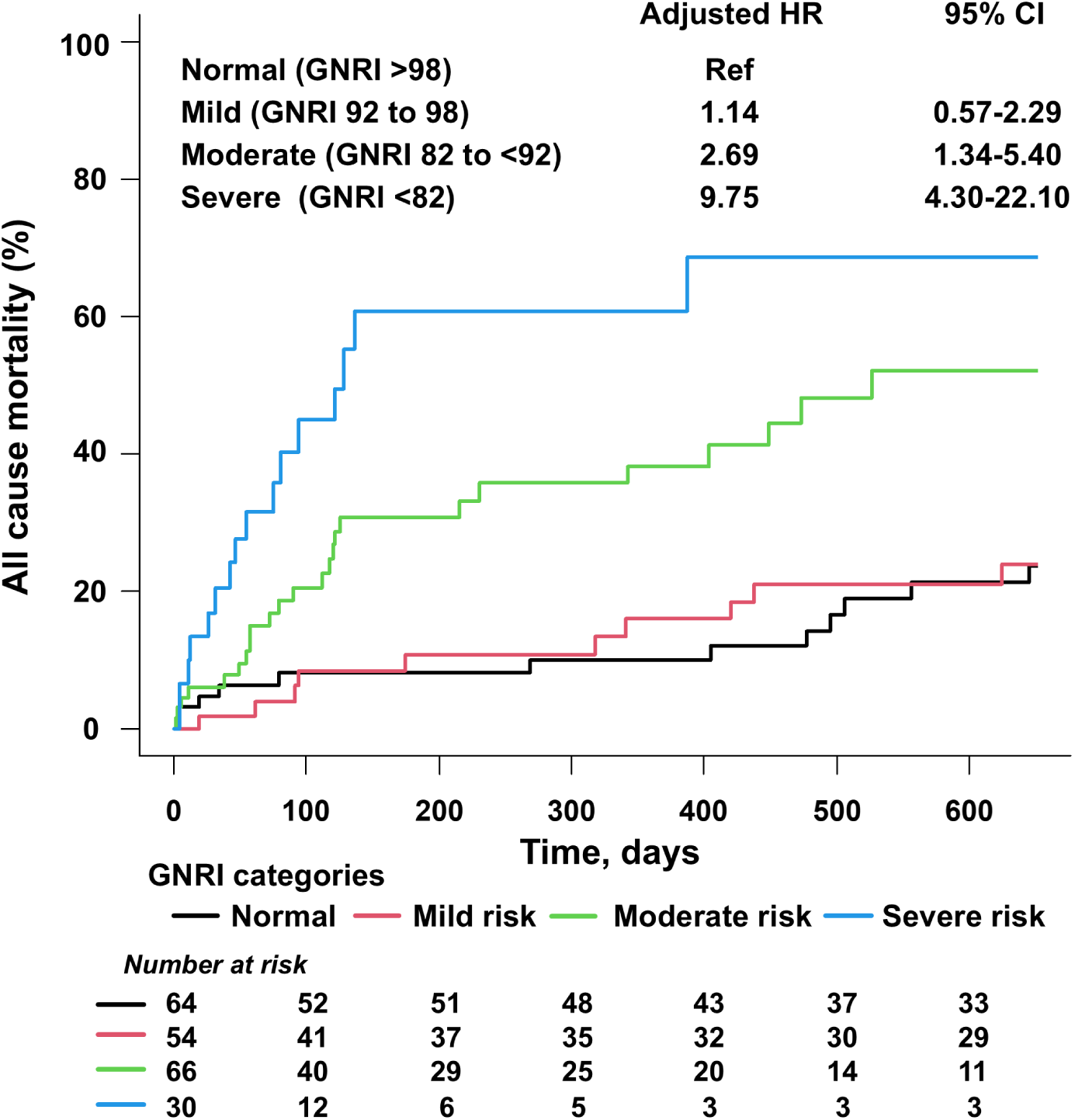
The GNRI risk categories and all-cause mortality. The moderate GNRI risk and severe GNRI risk were associated with higher rates of all-cause mortality after multiple adjustments. Adjusted HR of the GNRI risk was calculated by using a multivariable Cox hazard analysis with adjustments for age, sex, hypertension, diabetes mellitus, dyslipidemia, estimated GFR <60 mL/min/1.73m², and NYHA class. The normal GNRI category was used as a reference. Abbreviations: CI, confidence interval; HR, hazard ratio. Otherwise, as in Figure 1.

**Figure 3.**
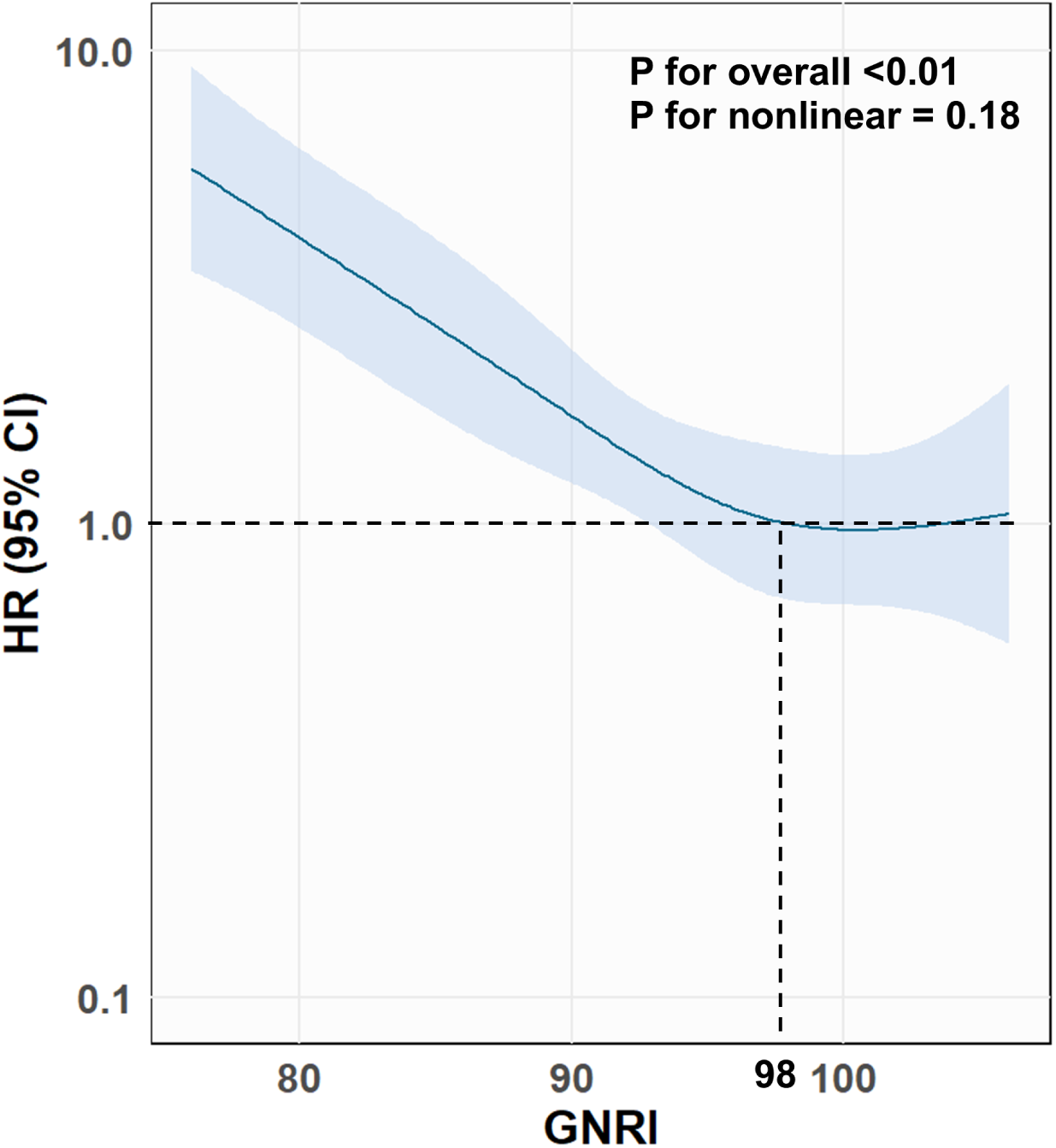
The GNRI in a continuous metric and all-cause mortality. The association between the GNRI and HR for all-cause mortality was demonstrated by a restricted spline curve (knot = 4, reference = GNRI 98). Age and sex were adjusted to calculate HR. Abbreviations as in Figures 1 and 2.

**Figure 4.**
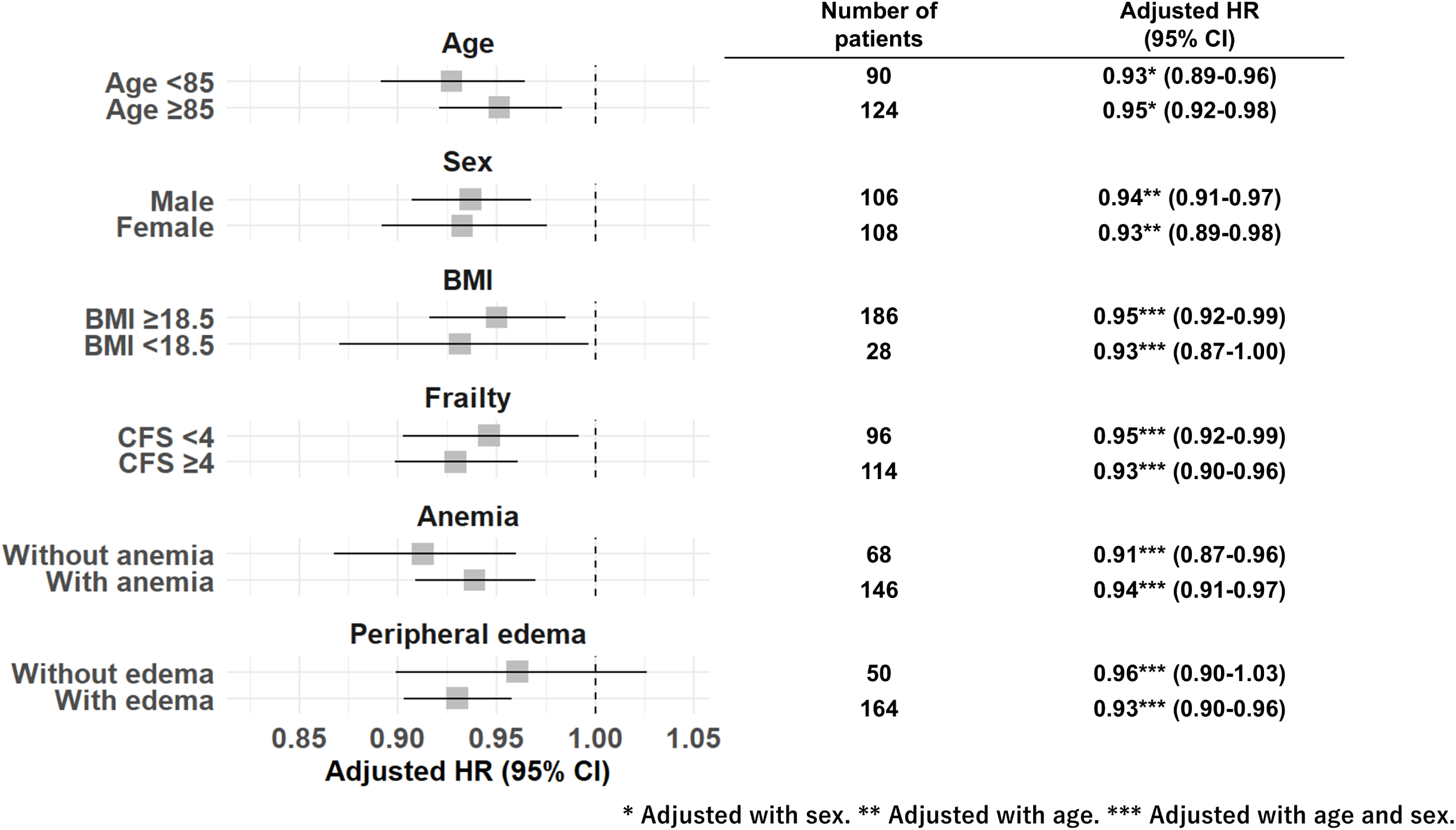
Subgroup analysis. The GNRI (as a continuous metric) showed consistent association with all-cause mortality regardless of age ≥85, sex, BMI <18.5, CFS ≥4, and anemia. HR was calculated with adjustments for age and sex. Abbreviations: BMI, body mass index; CFS, Clinical Frailty Scale. Otherwise, as in Figures 1 and 2.

**Table 2.** Multivariable Cox hazard analysis to investigate the association of GNRI with all-cause mortality.

|  | Crude |  |  | Adjustments (GNRI category) |  |  | Adjustments (continuous GNRI) |  |  |
| --- | --- | --- | --- | --- | --- | --- | --- | --- | --- |
|  | HR | 95% CI | P-value | HR | 95% CI | P-value | HR | 95% CI | P-value |
| Age | 1.08 | 1.04-1.12 | <0.01 | 1.07 | 1.03-1.11 | <0.01 | 1.06 | 1.02-1.10 | <0.01 |
| Female | 0.90 | 0.57-1.41 | 0.65 | 0.81 | 0.51-1.28 | 0.36 | 0.80 | 0.51-1.27 | 0.35 |
| Hypertension | 1.30 | 0.68-2.46 | 0.43 | 1.50 | 0.75-3.03 | 0.25 | 1.57 | 0.79-3.11 | 0.19 |
| Diabetes mellitus | 0.53 | 0.31-0.93 | 0.03 | 0.72 | 0.38-1.34 | 0.16 | 0.73 | 0.39-1.36 | 0.32 |
| Dyslipidemia | 0.46 | 0.24-0.88 | 0.02 | 0.60 | 0.30-1.20 | 0.22 | 0.67 | 0.34-1.34 | 0.26 |
| NYHA class |  |  |  |  |  |  |  |  |  |
| I/II | Ref |  |  | Ref |  |  | Ref |  |  |
| III | 0.68 | 0.33-1.38 | 0.28 | 0.74 | 0.35-1.57 | 0.30 | 0.70 | 0.33-1.47 | 0.35 |
| IV | 1.00 | 0.51-1.97 | 1.00 | 0.90 | 0.43-1.86 | 0.77 | 0.78 | 0.38-1.60 | 0.49 |
| Estimated GFR <60 mL/min/1.73m<sup>2</sup> | 1.37 | 0.82-2.28 | 0.23 | 1.70 | 0.99-2.95 | 0.06 | 1.52 | 0.89-2.61 | 0.13 |
| GNRI category |  |  |  |  |  |  |  |  |  |
| Normal | Ref |  |  | Ref |  |  |  |  |  |
| Mild risk | 1.44 | 0.74-2.79 | 0.28 | 1.14 | 0.57-2.29 | 0.71 |  |  |  |
| Moderate risk | 2.75 | 1.45-5.21 | <0.01 | 2.69 | 1.34-5.40 | 0.01 |  |  |  |
| Severe risk | 6.47 | 3.12-13.40 | <0.01 | 9.75 | 4.30-22.10 | <0.01 |  |  |  |
| GNRI (per 1 increment) | 0.94 | 0.92-0.96 | <0.01 |  |  |  | 0.92 | 0.90-0.95 | <0.01 |

## Discussion

We investigated the impact of the severity of malnutrition at admission on all-cause mortality of 214 older adults with AHF whose mean age was 85 years. Of those, a significant number of patients (14%) had severe nutritional risk (GNRI <82). Our results revealed a clear association between GNRI-based malnutrition severity and mortality risk, with an 8% decrease in mortality risk per 1-point GNRI increase, even in the oldest-old (those aged 85 years or older). Moreover, moderate or severe GNRI risk categories were independently associated with 2.7-fold or 9.8-fold increased HRs of all-cause mortality compared to the normal GNRI risk category, respectively. By evaluating not only the presence of malnutrition but also its severity, the mortality risk of older adults with AHF could be more accurately estimated (**Graphical abstract**).

## Severe malnutrition in older adults with AHF

In older adults with AHF, we revealed a high prevalence of malnutrition at admission, reaching 70% of at least mild nutritional risk (GNRI <98) and 45% of worse than moderate nutritional risk (GNRI <92). Malnutrition is common in older adults with heart diseases, such as atrial fibrillation (23), coronary artery disease (24), valvular heart disease (25, 26), HF with preserved ejection fraction (18), and AHF (8-15). Although the prevalence of malnutrition depends on the study population and the used assessment tool, AHF is one of the heart diseases in which malnutrition is frequently observed (8-10). According to previous reports from Japan (mean age, 77-79 years old), the prevalence of malnutrition (defined as GNRI <92) was 33-48% in patients with AHF (8, 12-14). This was comparable to our result (45%). However, in this study, we also demonstrated that 14% of patients had a severe nutritional risk (GNRI <82) at admission. Although the prevalence of severe malnutrition in AHF has been unclear, our results might provide insight into the importance of severe malnutrition in the oldest-old population. Aging plays a key role in the development of malnutrition (7, 27). Considering the aging trend of patients with AHF (4), the prevalence of severe malnutrition in patients with AHF will increase, and malnutrition in AHF will be a more critical concern. Furthermore, we showed a worsening trend in frailty as nutritional risk increased. In patients with HF, a complex interplay of neurohormonal derangement, inflammation, appetite suppression, malabsorption, and decreased mobility contributes to the development of malnutrition and muscle wasting (7, 28). These wasting conditions likely act as mutual aggravators, contributing to poor clinical outcomes. Therefore, in older adults with AHF, we should check for co-existing frailty when malnutrition is suspected.

## The prognostic impact of increasing malnutrition severity

We revealed an increasing risk of mortality with worsening of the GNRI in older adults with AHF. Previous studies reported the usefulness of the GNRI in estimating the mortality risk of older adults with AHF (8, 9, 12-15). Nakamura et al. reported that GNRI <92 was associated with all-cause mortality of AHF in patients aged 80 years or older (15). Their study population was the oldest (mean age: 87 years) among the previous studies (8, 9, 12-15). However, similar to other studies (8, 12-14), they dichotomized the study population by GNRI <92 as the definition of malnutrition (15). Therefore, the clinical importance of the severity of malnutrition in older adults with AHF (especially, in the oldest-old patients) remained unclear. In this study, we demonstrated that moderate nutritional risk (GNRI 82 to <92) and severe nutritional risk (GNRI <82) were associated with all-cause mortality, and there was an increasing trend in mortality with worsening GNRI. Furthermore, we confirmed the association of the GNRI with all-cause mortality in patients aged 85 years or older. Moreover, we performed multivariable analysis and subgroup analysis to adjust for comorbidities or vulnerable conditions related to elderly AHF, and they showed a consistent association between the GNRI and all-cause mortality. This highlighted the importance of the malnutrition severity to estimate the mortality of older adults with AHF appropriately and suggested the usefulness of the GNRI in the oldest-old patients (those aged 85 years or older).

## Clinical implication

We demonstrated a considerable 1-year mortality after AHF in patients with moderate GNRI risk (33%) and severe GNRI risk (50%), which was much higher than previously reported (11-27%) (2-6). As we demonstrated, worse nutritional status was associated with vulnerable conditions, including older age, underweight, frailty, and anemia. These suggest that the GNRI could enable us to screen high-risk, vulnerable patients requiring comprehensive care early after admission. In addition to the optimization of guideline-directed medical therapy for HF, support for coexisting comorbidity might contribute to better outcomes in malnourished patients with AHF. Regarding nutritional intervention for AHF, randomized controlled trials are sparse. Further, the causal relationship between malnutrition and the status of HF is difficult to discuss. However, a few trials demonstrate that nutritional intervention improves mortality, quality of life, and re-admission events of patients with HF (29, 30). These may support the importance of individualized nutritional treatments for older adults with AHF.

## Limitations

This study has several limitations. First, since this was a retrospective, single-center study, there could be a selection bias. However, this study, conducted in a rural area in Japan, could follow the aging population until the occurrence of the event. Second, although we calculated the GNRI at admission, volume congestion in AHF might influence the assessment of the nutritional status using the GNRI. Instead, we proposed the usefulness of the GNRI as a prognostic marker for older adults with AHF. The subgroup analysis showed a consistent association between GNRI and mortality in patients with peripheral edema, which might support this application. Third, due to the small number of events, we could not adjust for some confounding factors, such as LVEF and medical therapy, that might have affected the mortality. Finally, the temporal change in nutritional status was not assessed in this study. Future prospective studies will be needed.

## Conclusions

Moderate and severe GNRI risk categories and lower GNRI were independently associated with all-cause mortality in older adults with AHF. Malnutrition severity assessed by the GNRI could be useful in estimating the mortality risk for older adults with AHF, even in patients aged 85 years or older.

## Acknowledgments

The authors acknowledge the cardiology-unit staff in the Japanese Red Cross Ogawa Hospital for their technical support in this study.

## Data availability

All data are available from the corresponding author on reasonable request.

---

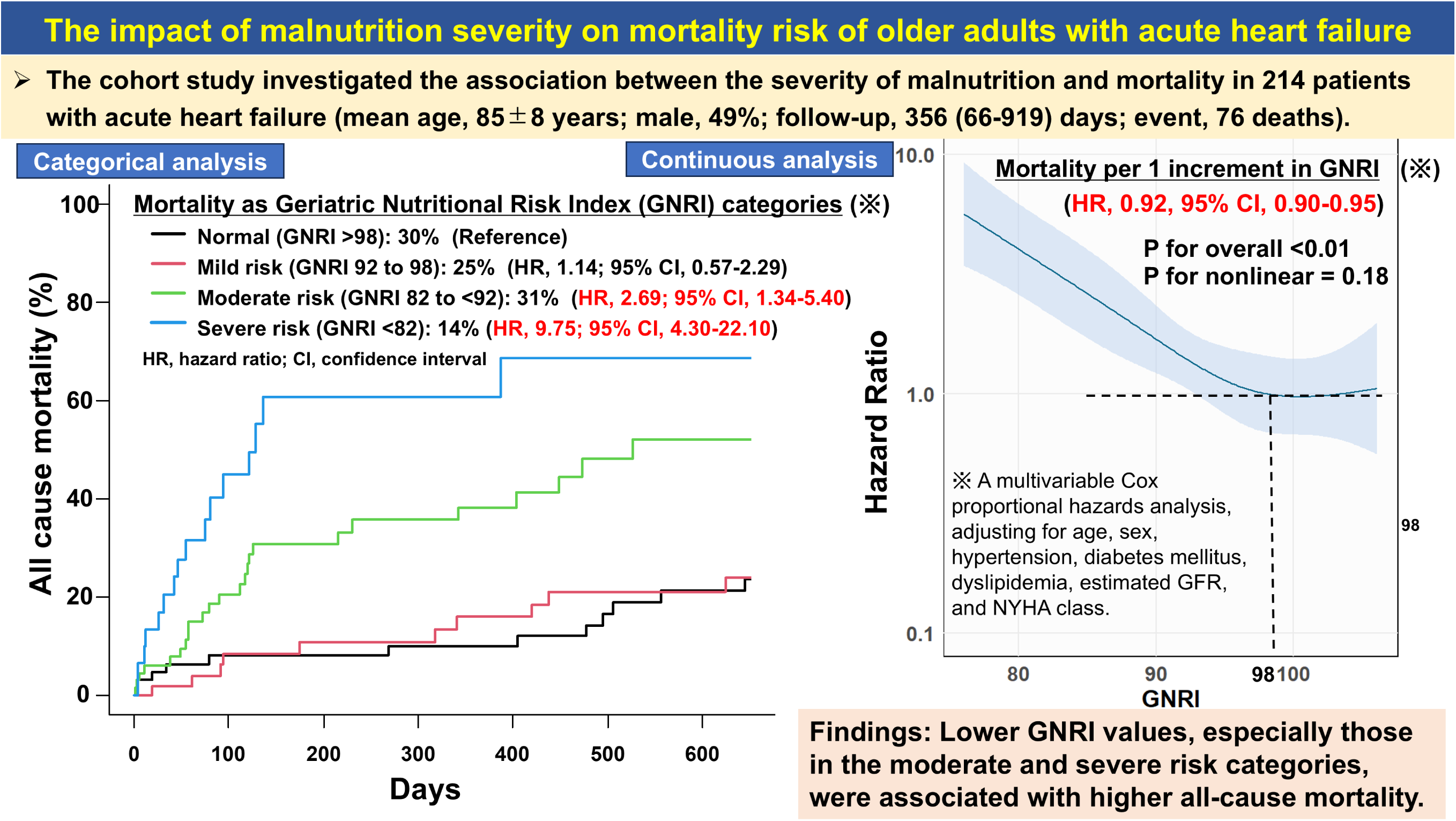

